# Post-Operative Bleeding After Facial Cleft Repair: A Retrospective Review of TriNetX Database

**DOI:** 10.1101/2024.08.13.24311878

**Authors:** Emily L. Isch, Aviana Duca, Theodore E. Habarth-Morales, D. Mitchell Self, Edward J. Caterson

**Affiliations:** Department of Surgery, Thomas Jefferson University Hospital, 1025 Walnut St, Suite 100, Philadelphia, PA, 19107, USA; Sidney Kimmel Medical College, Thomas Jefferson University, 1025 Walnut St, Philadelphia, PA, 19107, USA; Department of Neurosurgery, Thomas Jefferson University, 1025 Walnut St, Suite 100, Philadelphia, PA, 19107, USA; Department of Surgery, Division of Plastic Surgery, Nemours Children’s Health, Wilmington, DE, 19803, USA

**Author notes:** Corresponding author: Emily L. Isch, MD 840 Walnut Street 15th Floor Philadelphia, PA 19107. **Financial Disclosure Statement:** The authors have no financial disclosures to report.

**Keywords:** cleft palate, hemorrhage, post-operative complications

## Abstract

**Introduction:** The objective of this article is to assess and describe the incidence of postoperative hemorrhage following cleft palate surgery (palatoplasty), specifically focusing on need for return to the operating room for management of post-operative hemorrhage.

**Methods:** The TriNetX federated database was used to identify patients with a diagnosis of cleft lip and/or palate undergoing primary cleft palate repair over a twenty-year period from 2003 until2023. Primary endpoints assessed include post operative hemorrhage resulting in blood transfusion and/or return to the operating room; Kaplan-Meier analysis was used for statistical analysis.

**Results:** A total of 13,161 patients with cleft lip or palate over the last 20 years underwent operative intervention (palatoplasty). Of those patients, ninety-seven patients were found to have diagnosis of post-operative bleeding (confidence interval 1.196 +/-0.606). One hundred and seventy-five patients experienced post operative hemorrhage requiring transfusion of blood product (CI 1.491 +/-0.952). Seventy patients required return to the operating room for post-operative bleeding in the immediate post operative period.

**Conclusions:** Historic reporting of post-operative bleeding suggests a moderate rate of post-operative hemorrhage rate following palatoplasty, occasionally necessitating transfusion and return to operating room following index palatoplasty. Our retrospective review of a national database demonstrates a lesser incidence of post-operative hemorrhage than previously noted.

## Introduction

Cleft lip and palate represent some of the most prevalent congenital anomalies affecting the craniofacial region. Their occurrence may manifest as isolated deformities or part of broader syndromic spectrums, implicating both environmental and genetic etiologies. Within the United States, epidemiological data suggest that cleft palate alone manifests in approximately one in every 1,700 live births [1]. The embryological process—characterized by the fusion of the palate and lip at the midline between the sixth and ninth weeks of gestation—is intricate. A cleft palate typically arises from the failure of the medial nasal processes to merge with the maxillary processes, a critical step in forming the primary palate [2].

Interventions for children diagnosed with a cleft palate are not merely corrective but are a series of reconstructive procedures extending from infancy into early adulthood, aimed at addressing the multifaceted aesthetic and functional deficits concomitant with this anomaly. Palatoplasty, ideally performed between nine and eighteen months of age, is strategically timed to preempt the onset of speech while preserving the integrity of palatal functions and pressures [3]. Surgical methodologies for palatoplasty are diverse, tailored by institutional protocols and the surgeon’s expertise, yet unified by a triad of objectives: defect rectification, facilitation of speech development, and preemptive measures against maxillary growth perturbations that could precipitate dentoalveolar anomalies [4].

Despite advancements, palatoplasty is beset with potential complications encompassing wound dehiscence, infection, scarring, hematoma, respiratory difficulties due to upper airway manipulation, and adverse reactions to anesthesia. Additionally, postoperative speech complications and velopharyngeal dysfunction are of considerable concern. Notably, postoperative hemorrhage remains a critical issue, carrying a significant risk of morbidity and, in severe cases, mortality if not promptly managed [5].

The literature within plastic surgery underscores the significance of meticulous hemostasis and vigilant postoperative monitoring to circumvent hemorrhagic complications. Immediate postoperative care, typically within the first 24 hours, is crucial for detecting signs of airway compromise or active bleeding, which may necessitate rapid medical intervention. Discharge protocols hinge on the assurance of stable hemostasis, adequate feeding, and tolerability of prescribed medications, generally between the first and third postoperative days. The Pediatric Perioperative Cardiac Arrest Study (POCA) identifies hemorrhagic hypovolemia as a leading cause of perioperative mortality, emphasizing the exigency of standardized postoperative regimens adherent to evidence-based practices. This study endeavors to quantify the incidence of postoperative hemorrhage following palatoplasty in a pediatric cohort, delineating the clinical recourse to transfusion requirements and surgical re-interventions. Utilizing the TriNetX federated database, we conducted a retrospective analysis spanning two decades (2003-2023) to assess the prevalence of postoperative hemorrhage and its management in patients treated for cleft lip and/or palate.

## Methods

This investigation harnessed the expansive potential of the TriNetX national database, an aggregated compendium of electronic health records and clinical data amalgamated from a multitude of healthcare institutions. The breadth of this dataset permits a comprehensive and multifaceted approach to retrospective analyses within the field of medical research. Utilizing this resource, we meticulously identified a cohort of patients diagnosed with cleft lip and/or palate who underwent palatoplasty within the designated study period extending from 2003 to 2023. The inclusion criteria were strictly adhered to, ensuring a robust and relevant patient population for analysis.

We delineated our primary endpoints to encompass instances of postoperative hemorrhage with the consequent necessity for either blood transfusion or surgical re-intervention, indicative of severe hemorrhagic events. Such clinical endpoints serve as critical markers for assessing the quality and outcomes of surgical interventions.

Employing a Kaplan-Meier survival analysis, we sought to elucidate the time-to-event data for our population, offering insights into the longitudinal risk profile for postoperative hemorrhage within this demographic. This statistical method provided a probability distribution of time until the occurrence of hemorrhage, allowing for an estimation of the survival function and the median survival time.

The analytical rigor applied in this methodological approach ensures a high caliber of data integrity and reliability of results, suitable for contributing to evidence-based practices in the field of pediatric plastic surgery.

## Results

This longitudinal analysis, spanning a period of two decades from 2003 to 2023, encompassed 13,161 patients who underwent surgical intervention for cleft lip or palate anomalies. Our examination yielded that 1.3% of this population, corresponding to 175 patients, encountered postoperative hemorrhage severe enough to warrant transfusion of blood products, with a confidence interval (CI) of 1.491 +/-0.952. Within this subset, a smaller proportion, accounting for 0.7% or 97 patients, were identified with clinically significant postoperative bleeding, denoted by a CI of 1.196 +/-0.606.

Crucially, 0.5% of the patient cohort, translating to 70 individuals, necessitated re-operative intervention in the immediate postoperative timeframe due to hemorrhagic complications. This subset of patients required operative management, underscoring the critical nature of surgical oversight in the immediate recovery phase.

These empirical findings elucidate the incidence and necessitate a nuanced understanding of postoperative hemorrhage within the domain of palatoplasty. The data derived from this study augment the extant corpus of medical knowledge regarding the post-surgical trajectory of patients with cleft palate anomalies, with implications for both surgical practice and postoperative care protocols.

## Discussion/Conclusion

Postoperative hemorrhage in the context of palatoplasty represents a clinical challenge that, while not exhaustively documented in current literature, is acknowledged as a non-trivial complication with incidences reported between approximately 1.8% to 5.5%. The variance in documented blood loss, which can be as minimal as 16 milliliters, underscores the influence of patient-specific pathophysiology, the surgeon’s acumen, and the chosen surgical technique on postoperative outcomes [6].

The present retrospective analysis encompassed 366 individuals who underwent primary palatoplasty. Within this subset, the incidence of significant postoperative hemorrhage necessitating medical or surgical intervention was notably lower than extant literature suggests— 1.6% experienced late bleeding, and only 0.7% encountered events severe enough to warrant readmission for transfusion or surgical hemostasis. These figures offer a substantial deviation from the higher rates historically reported and may reflect improvements in surgical techniques or postoperative care protocols [7].

Our findings are further contextualized by a comparative analysis with a prospective cohort where 2.4% required operative management for hemorrhage in the immediate postoperative phase. The 0.5% of our cohort requiring similar intervention signifies an appreciable reduction, albeit within a different study design. Additionally, the 1.3% transfusion rate observed in our study prompts a discussion on regional healthcare disparities and the continuity of care post-discharge, which may influence the management of postoperative complications.

The evidence marshaled herein underlines the imperative for an in-depth understanding of postoperative hemorrhage etiologies to mitigate such risks and enhance patient outcomes. Future research directions should aim to dissect the nuances of early versus late bleeding episodes, explore surgical techniques that have proven efficacious in hemostasis, and establish a set of criteria for operative revision. Moreover, the potential role of tranexamic acid (TXA) in ameliorating perioperative hemorrhage warrants systematic investigation to inform evidence-based clinical guidelines [8].

In summation, the elucidation of optimal postoperative management strategies and the refinement of surgical interventions remain paramount. This study contributes to a burgeoning body of evidence poised to inform both the operative and postoperative paradigms, ultimately advancing the standard of care for patients afflicted with cleft palate anomalies.

## Data Availability

All data produced in the present work are contained in the manuscript

## References

1. “Facts about Cleft Lip and Cleft Palate.” Centers for Disease Control and Prevention, Centers for Disease Control and Prevention, 28 June 2023, https://www.cdc.gov/ncbddd/birthdefects/cleftlip.html#:~:text=How%20Many%20Babies%20are%20Born,palate%20in%20the%20United%20States.

2. Yap, Joshua. “Cleft Lip and Palate: Radiology Reference Article.” Radiopaedia, Radiopaedia.org, 8 May 2023, radiopaedia.org/articles/cleft-lip-and-palate?lang=us.

3. Skolnick GB, Keller MR, Baughman EJ, Nguyen DC, Nickel KB, Naidoo SD, Olsen MA, Patel KB. Timing of Cleft Palate Repair in Patients With and Without Robin Sequence. J Craniofac Surg. 2021 May 1;32(3):931–935. doi: 10.1097/SCS.0000000000007311. PMID: 33290333; PMCID: PMC8680105.

4. Agrawal, Karoon. “Cleft Palate Repair and Variations.” Indian Journal of Plastic Surgery : Official Publication of the Association of Plastic Surgeons of India, U.S. National Library of Medicine, Oct. 2009, https://www.ncbi.nlm.nih.gov/pmc/articles/PMC2825076/.

5. Denning, S, et al. “Anaesthesia for Cleft Lip and Palate Surgery.” BJA Education, U.S. National Library of Medicine, Oct. 2021, https://www.ncbi.nlm.nih.gov/pmc/articles/PMC8446239/.

6. Mortada, Hatan, et al. “Is tranexamic acid effective in minimizing blood loss in cleft palate repair? A rigorous assessment through a comprehensive systematic review and meta-analysis.” British Journal of Oral and Maxillofacial Surgery, Jan. 2024, 10.1016/j.bjoms.2023.12.019.

7. Wheeler JS, Sanders M. Late Bleeding Following Cleft Palate Repair: An Under-Reported Finding? J Craniofac Surg. 2022 Mar-Apr 01;33(2):607–609. doi: 10.1097/SCS.0000000000008135. PMID: 34519712.

8. Bharadwaj A, Khandelwal M, Bhargava SK. Perioperative neonatal and paediatric blood transfusion. Indian J Anaesth. 2014 Sep;58(5):652–7. doi: 10.4103/0019-5049.144679. PMID: 25535431; PMCID: PMC4260315.

